# Time to initiate trophic feeding and predictors among preterm neonates admitted at General Hospitals in Tigray, 2025

**DOI:** 10.1101/2025.10.13.25337936

**Authors:** Teklebrhan Welderufael Kidane, Zeray Baraki, Asefa Iyasu, Tekle Gebremeskel Ygzaw, Binyam Gebrehiwet Tesfay, Ngsti Gebremichael Beyene, Teklewoini Mariye Zemicheal, Nebiat Desale Gidey, Geberziher Gebreslassie Welearegay

## Abstract

**Back ground:** Trophic feeding generally refers to providing small quantities of enteral feeding soon after birth. As a result this study is aimed to assess time to initiation of TF and its predictors among preterm neonate in Tigray region.

**Method:** A prospective, institutional-based follow-up study was conducted on 193 preterm neonates admitted to the Neonatal intensive care unit, with participants’ selected using systematic random sampling from a group of public hospitals. Data collection period was from December 20, 2024 to February 30, 2025. Data was entered into Epi data version 4.7 and then exported to STATA version 14 for cleaning and analysis. To compare survival curves, Kaplan-Meier analysis and the log-rank test was used, bivariate and multivariate Cox regression analysis were used, all statistical tests was considered significant at a p-value of <0.05.

**Result:** A total of 193 neonates were followed for 8382 person-hours of risk time and 173 (89.6%) of neonates were initiated trophic feeding. The incidence rate of initiating trophic feeding was 2 per 100 person hours’ observations with a median time of 45 hours (95% CI: 42-56). Birth weight <1500 gram (AHR: 0.16,95% CI:0.075-0.35), APGAR score at first minute < 7 (AHR: 0.46,95% CI:0.26-0.76),APGAR score at fifth minute < 7 (AHR: 0.38,95%CI :0.21-0.68,, having respiratory distress syndrome (AHR: 0.41,95% CI:0.25-0.66, and absence of Kangaroo mother care (AHR: 0.41,95% CI:0.21-0.77), were statistically Significant associated factors for the delay of initiation of trophic feeding.

**Conclusion:** In this study, a significant delay in the initiation time of trophic feeding and several predictors were identified. Therefore, health institutions should work on these predictors to shorten the initiation time and reduce complications associated with the delay.

## Introduction

Trophic feeding (TF) is typically involves administering small amounts of enteral feeding (EF) shortly after birth with amount (1–2 mL/kg per dose or 10-15 mL/kg per day) to stimulate and supply essential nutrients to the developing gastrointestinal tract; this minimal enteral nutrition (MEN) helps promote intestinal development, improve feeding tolerance, prepare the gut, and provide hypo-caloric nourishment to support the immature digestive systems of preterm neonates. (1, 2).

Preterm birth is defined as birth before 37 completed weeks of gestation or fewer than 259 days from the first date of a woman’s last menstrual period (3, 4).These infants experience a nutritional emergency as they are suddenly removed from nutrient rich environment in-utero to the extra uterine life where nutrition is harder to initiate and later on maintain (5).

In the neonatal period, proper nutritional care is crucial for minimizing the risk of both short- and long-term negative outcomes. It ensures that the baby receives the necessary nutrients to match the growth rate and body composition of a healthy fetus at the same gestational age (GA) and this includes aspects such as weight, length, head circumference, organ size, tissue composition (including cell number and structure), as well as blood and tissue nutrient levels, and overall developmental progress (6, 7).

The World Health Organization (WHO) 2021 strongly recommends, based on moderate-certainty evidence, that enteral feeding through a nasogastric tube should be started as early as possible, ideally within the first day after birth, for preterm or low birth weight (LBW) infants, including those weighing less than 1.5 kg or born before 32 weeks of gestation and this recommendation is supported by evidence showing moderate benefits, including reduced mortality and shorter hospital stays (8).

The two classification with recommendation of TF feeding practice according to the Ethiopian Neonatal Intensive Care Unit (NICU) guide line 2021 in preterm neonates are early initiation of TF start within 24 hours of birth and delayed initiation of TF which start after 24 hours of birth (9).

Delayed initiation of TF is a global concern in NICUs and has been identified as a significant independent predictor of poor growth among preterm neonates (10). Globally complications arising from preterm birth are the leading cause of neonatal deaths accounting for 35% of the 3.1 million neonatal deaths each year with 15 million preterm births globally those born before 32 weeks of gestation are at the highest risk of illness and death and nearly half of all under-five deaths occur among neonates (infants under 28 days old) (11).Similarly in Ethiopia preterm birth complications continue to be a major cause of neonatal mortality and despite global efforts to reduce these complications the rates of preterm birth remain high with significant delays in interventions such as early trophic feeding (11–13).

Some of the inherent that delay to initiate TF were Increased incidence of necrotizing enterocolitis NEC, prolonged hospital stays, and higher mortality rates (10, 14).

TF is a core and widely acknowledged approach that aids in reducing the risk of complications linked to enteral fasting, including necrotizing enterocolitis (NEC), infections, prolonged hospital stay, metabolic problems, and postnatal growth failure (PGF) in preterm neonates (14–21).

Delayed start of TF in preterm neonates can lead to inadequate growth, heightening the chances of nutritional deficiencies, hindered brain development, sepsis, feeding difficulties, low weight, and reduced survival rates among preterm neonates (10, 13, 22, 23).Similarly, late enteral feeding and long hospital stay of preterm infants leads to, placing a significant economic burden on families and healthcare service system, stressful hospital stays, neonatal intensive care expenditures, ongoing health care, and educational demands (24, 25).

Preterm birth is the primary cause of neonatal and under-5 mortality worldwide, and the associated healthcare burdens and nutrition-related complications are unsustainable, especially in resource-constrained areas (26).Every year, there are 15 million preterm births globally, with infants born before 32 weeks of gestation facing the greatest risk of morbidity and mortality from this undernutrition in these preterm infants is linked to severe outcomes, including a higher risk of death (11).

International studies have found that 80 to 90% of these newborns did not begin TF until they were 48 hours old which indicated a significant delay in the start of enteral feeding for preterm infants (14). In Africa, only a small percentage of infants begin minimal feeding early, resulting in significant consequences. For instance, a study conducted in Uganda found that starting TF more than 48 hours after birth was linked to postnatal growth failure compared to initiating feeding sooner (5, 10). In addition study in Zambia reviled that neonate delayed enteral feeding cause 66% death compared with 6% in the early feeders (27).

Similarly, research from Ethiopia found considerable delays in TF for neonates, despite guidelines recommending that it be initiated within 24 hours of birth, 80–90% not starting feeding within 48 hours; furthermore, only 20% of infants received TF within 24 hours, while 29% did not survive to discharge, and 86.2% experienced extra uterine growth restriction (10, 28).(29).

Several factors were recognized as major predictors of this delay, including an APGAR score below 7 at one minute, APGAR score less than seven at five minute, GA under 34 weeks, respiratory distress syndrome (RDS),Very low birth weight, hemodynamic instability, perinatal asphyxia (PNA), cesarean section (CS) delivery, Absence of KMC service sepsis, hypothermia and being born out of the study hospitals (10, 13, 14, 28).

Since there is limited evidence regarding time to initiate TF among preterm neonates admitted to NICU on prospective follow up is not adequate searched and have limited data in Ethiopia especially recently in Tigray, further research is needed for further investigation the problem. So, it is important to investigate the factors that delay the initiation of TF for gaining a deeper understanding, enhance the health quality of preterm neonates and reducing healthcare costs and improving long-term outcomes for families.

Therefore, the problem needs investigation by understanding the predictors of TF initiation is crucial for successful modification, development, and implementation of appropriate feeding guidelines for premature neonates and improving clinical practice. As a result this study is aimed to assess time to initiation of TF and its associated factor of preterm neonate in Tigray region.

Finally, the findings of this research will serve as a crucial resource for stakeholders in neonatal care.

## Methods and materials

### Study area and period

The study was carried out in six General hospitals the Tigray, Ethiopia, from August 1, 2024, to May 30, 2025. The region has 2 specialized hospitals, 14 general hospitals, 24 primary hospitals, 230 health centers, and 741 health posts. All the general hospitals are organized into different service areas, including term, preterm, isolation, and procedure rooms with KMC and maternal waiting rooms. Each NICU ward has around 10-16 healthcare providers. There are two specialized hospitals in the region, and both of them have separate admission rooms like KMC service rooms, and each of them has 20-40 healthcare workers. Data collection period was carried out from December 20, 2024 to February 30, 2025.

### Study design

Institution based, prospective follow up study design was conducted.

### Source of population

All preterm neonates (< 37 weeks GA) admitted to NICU during the study period at public general hospitals in, Tigray, Ethiopia.

### Study population

All preterm neonates (< 37 weeks GA) admitted to NICU during the study period at the selected public general hospitals.

### Eligibility criteria

#### Inclusion criteria

Preterm neonates (less than 37 weeks GA) admitted to the neonatal intensive care unit (NICU) of the study general hospitals during the study period.

#### Exclusion criteria

Preterm contraindicated for feeding like pre-diagnosed Stage II or III necrotizing enterocolitis and Stage III asphyxia. Neonates with significant gastrointestinal malformations, such as esophageal atresia, imperforate anus, duodenal and jejunal atresia, intestinal obstruction or perforation, and paralytic ileus.

Neonates with, unknown APGAR scores, gestational ages and home deliveries were excluded from the study.

### Sample Size Determination

Sample size was calculated using Stata version 14.2 by considering AHR of 0.63,probability of event (neonate initiated TF) was 0.85 (28), two-sided, 5% significant level, power of 80% and 10% probability of withdrawing from a study. After checking all significant factors from the previous study the maximum sample size obtained was 193. The total sample size was proportionally allocated to each study hospitals.

### Sampling procedure

The study area was chosen through a simple random sampling method using a lottery approach. The hospitals included in the study are Aksum St. Mary Hospital, Shire Suhul Hospital, Adigrat Hospital, Maychew Hospital, Miyani Sheraro Hospital, and Adwa Hospital.

The total number of preterm neonates admitted to the NICU ward in the past two months was reviewed in each of the selected general hospitals, and a total of 66, 70, 64, 55, 60, and 60 admissions were obtained from Aksum St. Mary Hospital, Shire Suhul Hospital, Adigrat Hospital, Maychew Hospital, Miyani Sheraro Hospital, and Adwa Hospital, respectively.

Preterm neonates admitted to the NICU ward from December 20, 2023 to February 30, 2024 from the six selected public general hospitals totaled 375. The samples were allocated proportionally to each hospital (34 from Aksum St. Mary Hospital, 36 from Shire Suhul Hospital, 33 from Adigrat Hospital, 28 from Maychew Hospital, 31 from Miyani Sheraro Hospital, and 31 from Adwa Hospital). Finally, the sample size was 193.

After allocation, finally, a systematic random sampling method (K = N/n = 375/193 ≈ 1.9 ≈ 2) was used. The first participant was selected randomly, and then every Kth interval (every 1^st^ interval) was chosen as a study participant during admission.

### Study variable

#### Dependent Variable

Time to initiate TF

#### Independent Variable

**Maternal sociodemographic factors** such as age of the mother, residence, educational status and maternal obstetric and medical related factors, mode of delivery and maternal HIV AIDS status.

**Neonatal related factors**; birth weight, sex, gestational age, first minute Apgar score, fifth minutes Apgar score, perinatal asphyxia, sepsis, hemodynamic instabilities, respiratory distress syndrome weight for gestational age and hypothermia,

**Health service-related factors**; place of delivery and KMC service.

### Operational Definitions

**TF**: The first minimal enteral feeding administering small volumes of enteral feeds (typically 10– 20 mL/kg/day) to stimulate the development of the immature gastrointestinal tract in preterm given to stimulate the gut (30).

**Early initiation of TF**: Preterm neonates start TF within 24 hours of birth (28).

**Delayed initiation of TF**: Preterm neonates start TF after 24 hours of birth (28).

**Survival time**: the length of time in hours followed starting from birth to the first TF (10).

**Event:** the preterm neonates who had started first TF within the follow-up period (10).

**Censored**: Preterm neonates who died, left against medical advice, transferred or referred before starting TF, or not started at end follow-up (14).

**Follow up time**: time from birth up to either the study subjects start TF or censored (10).

**Hemodynamic instabilities**: Blood group and RH incompatibility, anemia, polycythemia, bleeding disorders, blood glucose disturbances (10).

### Data Collection Tools and Procedure

Data collection was collected using a semi-structured, pretested questionnaire specifically designed to ensure both reliability and relevance. This instrument encompassed a comprehensive range of topics, including health service utilization, key determinants of maternal and neonatal health, and core sociodemographic variables. The questionnaire’s content was thoughtfully adapted and synthesized from a diverse body of established literature, ensuring it captured contextually significant and evidence-based insights relevant to the study objectives (10, 14, 28, 30).

Through interviews, chart reviews with prospective follow-up, and six nurses were involved in the data collection supervised by two supervisors. Data collectors (nurses working out of the institutions) and two supervisors were trained on how to follow up and itemize the included questionnaire, how to obtain client consent, and how to collect data accordingly. The supervisor reviewed the questionnaire for completeness.

### Data Quality Assurance

Before the actual data collection the questionnaire were adapted first in English, then translated to the local language Tigrigna, and then back to English to ensure consistency. Data collectors received two days training about the study and the content of the instrument in order to familiar about each question and follow up and it was also a mechanism of minimizing bias during the process of data collection.

During data collection, supervisors checked data for its completeness and missing information at each point. A pretest was done at Wukro K/awlaelo general hospital on 5% sample size.

### Data processing and Analysis

Data were entered into Epi Data Manager V 4.6 after being checked for completeness and consistency. It was subsequently exported to STATA version 14.1 for cleaning, coding, and analysis. During this process, the level of missing values, the presence of multicollinearity were reviewed, and the value was 2.01, along with the Cox proportional hazards regression model assumptions were checked using the Schoenfeld residual test, which yielded a value of 0.12, greater than 0.05, indicating that the assumptions were fulfilled.

To estimate survival time and cumulative probabilities of initiating TF, descriptive statistics, Kaplan-Meier survival curves, life tables, and hazard functions were applied. Additionally, the log-rank test was used to compare the survival curves of different categories. Finally, bi-variable cox regression analysis were conducted and -value of < 0.25 were used as a cut-off point to enter variables to multivariable Cox-regression and, a test was consider significant at P. value < 0.05.

### Dissemination of the result

The findings of this research will be submitted and presented to the College of Health Sciences at Axum University. Additionally, the results will be shared with general hospitals and the Tigray Regional Health Bureau (TRHB).Finally, Efforts will also be made to publish the study in a peer-reviewed scientific journal.

## Result

### Socio-demographic, clinical characteristics of mothers and neonates factors

A total of 193 preterm neonates were included in the final analysis with a 100 % response rate, of which 174 (87.6%) were started on TF, 20 (10.4%) were censored. One hundred twenty one (62.7%) and 72 (37.3%) of mothers were from urban and rural areas, respectively. The mothers were between the ages of 18 and 38, with a mean age of 25.72 years. Of them, 77 (39.9%) had finished secondary school. From the preterm neonates, 120 (62.2%) were GA >=34 weeks and all were low birth weight with a mean weight of 2300 gram ranging from 1000 to 24800 gram.

Fifty-six (29%) and 137(71%) neonates were males and females, respectively. Most of the births were spontaneous vaginal delivery167 (86.5%). Concerning weight for gestational age, out 159 (82.4%) were appropriate for their gestational age. The preterm neonates had a mean APGAR-score of 6.9 in the first minute and 7.4 in the fifth minute.

One hundred sixty (82.9%) of neonates had sepsis, 146 (75.6%) had hypothermia, and 88 (45.4%) of them did not receive KMC. Almost more than half 57.5%) of the neonates had a first-minute APGAR score of less than 7. Additionally, 65 (33.7%) and 59 (30.6%) of admitted neonates were had RDS and fifth minute APGAR score of less than 7.

**Table 1.** socio demographic and clinical characteristics of mothers and neonates to NICU in public general hospitals of Tigray Ethiopia 2024/25

| Socio-demographic,<br>health care and<br>clinical<br>characteristics | Category | Status |  | Total |
| --- | --- | --- | --- | --- |
|  |  | Start TF | censored |  |
| Maternal age | 18-24 | 106(93.8%) | 7(6.1%) | 113 |
|  | 25-34 | 58(80.6%) | 6(9.4%) | 64 |
|  | >=35 | 9(76.2%) | 7(43.8%) | 16 |
| Maternal education | Illiterate | 16(76.2%) | 5(23.8%) | 21 |
|  | Primary | 62(92.5%) | 5(7.5%) | 67 |
|  | Secondary | 72(93.5%) | 5(6.5%) | 77 |
|  | College and above | 23(82.1%) | 5(17.9%) | 28 |
| Residence | Urban | 108 (89.2%) | 13(10.8%) | 121 |
|  | Rural | 65 (90.3%) | 7(9.7%) | 72 |
| Birth weight in gram | 1500-2499gm | 149(92.5%) | 12(7.5%) | 161 |
|  | <1500 gm | 24(75%) | 8(25%) | 32 |
| Gestational age in<br>weeks | >=34 | 111(92.5%) | 9(7.5%) | 120 |
|  | <33 | 62(84.9%) | 11(15.1%) | 73 |
| 1st minute APGAR<br>score | <7 | 96(86.4%) | 15(13.5%) | 111 |
|  | >=7 | 77(93.9%) | 5(6.1%) | 83 |
| 5th minute APGAR score | <7 | 47(79.7%) | 12(20.3%) | 59 |
|  | >=7 | 126(94%) | 8(6%) | 134 |
| Sex | Male | 51(91.1%) | 5(8.9%) | 56 |
|  | Female | 122(91%) | 15(9%) | 137 |
| Perinatal Asphyxia | Yes | 69(88.5%) | 9(11.5%) | 78 |
|  | No | 104(90.4%) | 11(9.6%) | 115 |
| Hemodynamic instability | Yes | 51(78.5%) | 14(21.5%) | 65 |
|  | No | 122(95.3%) | 6(4.7%) | 128 |
| Respiratory Distress Syndrome | Yes | 50(76.9%) | 15(23.1%) | 65 |
|  | No | 123(96.8%) | 5(3.2%) | 127 |
| Weight for Gestational Age (GA) | AFGA | 144(90.5%) | 15(9.5%) | 159 |
|  | SFGA | 29(85.3%) | 5(14.7%) | 34 |
| Mode delivery | SVD | 154(92.2%) | 13(7.8%) | 167 |
|  | CS | 19(73.1%) | 7(26.9%) | 26 |
| Place of delivery | In born | 115(93.5%) | 8(6.5%) | 123 |
|  | Out born | 64(90.1%) | 7(9.9%) | 71 |
| Sepsis | Yes | 145(90.6%) | 15(9.4%) | 160 |
|  | No | 28(84.8%) | 5(15.2%) | 33 |
| Hypothermia | Yes | 132(90.4%) | 14(9.6%) | 146 |
|  | No | 41(88.2%) | 6(12.8%) | 47 |
| Kangaroo mother care (KMC) | Yes | 98(93.3%) | 7(6.7%) | 105 |
|  | No | 75(85.2%) | 13(14.8%) | 88 |

### Survival status of neonates on time to initiate TF

A total of 8382 person-hours of risk time were followed for 193 pairs of newborns and mothers. The follow-up period ranged from 12 hours to 84 hours. Out of 193 preterm newborns, 173 (8 95% CI: 89.67–89.73) were starting TF, and the remaining 20 (10.4%) were censored.

The median time to initiate TF for the entire follow up study was 45 hours (95% CI: 42–56) hours (interquartile range: 24-65) and the median follow up time was 43 hours (interquartile range: 24-60).

In the first day (24 hours), only 58 (30%) preterm neonates initiated TF and the remained 116 (58.5%) and 166 (86 %) started by the end of 48 and 72 hours. finally, only 8 neonates remained, with 7 starting TF, 1 being censored

The cumulative survival probabilities of preterm neonates by the end of 24, 48, 72 and 84 hours were 69.7% (95% CI: 0.6267 - 0.7569), 39.6% (95% CI: 0.3256 0.4648), 8% (95% CI: 0.0458 - 0.1273), 0.5% (95% CI: 0.0002 - 0.0450) respectively.

**Table 2.** Life table analysis of time to initiate trophic feeding among preterm neonates Admitted in NICU in General public Hospitals from December 20, 2024 to February 30, 2025.Tigray, Ethiopia.

| Interval | Beginning total | Start TF | Censored | Survival probability | 95% confidence interval |
| --- | --- | --- | --- | --- | --- |
| 0-24 | 193 | 58 | 5 | 0.6971 | 0.6267 - 0.7569 |
| 24-48 | 130 | 55 | 3 | 0.3957 | 0.3256 - 0.4648 |
| 48-72 | 72 | 53 | 11 | 0.0803 | 0.0458 - 0.1273 |
| 72-96 | 8 | 7 | 1 | 0.0054 | 0.0002 - 0.0450 |

### Comparison of survival among different categorical variables

Kaplan-Meier curves constructed to compare the overall survival patterns over time for between different groups. Similarly, the log rank test analysis indicated that there were substantial differences in median time to initiate TF among preterm neonates across categories of several variables..

Accordingly, there was a significant difference in median time to initiate TF: for BW 1500-2499 gram was 36 hours (95% CI: 33-39) compared to BW less than 1500 grams. 72 hours with (71-73) was statistically significant (log-rank test = 60, P < 0.001). Neonates with GA >= 34 weeks exhibited significantly earlier onset at 27 hours (95% CI: 25-29) compared to 60 hours (95% CI: 53-67) for those with GA < 34 weeks. Concerning clinical-related factors of preterm neonates, those who had RDS experienced a significantly delayed TF initiation of 70 hours (95% CI, 68–72) compared to those without RDS 27 hours (95% CI, 24–30), and this was statistically significant (log-rank test = 86.6, P < 0.001.

Similarly, the median time to initiate TF was significantly different for neonates with a first-minute APGAR score < 7, at 60 hours (95% CI, 57–63), compared to first-minute APGAR score >=7, at 24 hours (23.8–24.2) at P < 0.001 In a similar vein, neonates with a fifth-minute APGAR score < 7 minutes had 72 hours of median time to start TF at (95% CI, 71–73) compared to fifth-minute APGAR score >= 7 minutes at 30 hours (95% CI: 25–35) with p < 0.001). Finally, neonates without KMC had late commencement of TF, with a 65-hour median time to start TF (95% CI: 61-69) compared to neonates with KMC, 24 hours (95% CI: 24 (23.7-24.3)), which was statistically significant at P < 0.001.

**Table 3.** survival time and log rank analysis of preterm neonates according to different characteristics of neonates admitted to public General hospitals in NICU from December 20, 2024 to February 30, 2025, in Tigray, Ethiopia.

| covariates | category | median survival time in<br>hours<br>(95% ci) | log rank<br>(x2<br>value) | p-value |
| --- | --- | --- | --- | --- |
| Birth weight in gram | 1500-2499 | 36(33-39) | 60 | <0.001 |
|  | <1500 | 72(71-73) |  |  |
| Gestational age in<br>weeks | >=34 | 27(25-30) | 51.9 | <0.001 |
|  | <34 | 60(53-67) |  |  |
| 1st minute APGAR<br>score | <7 | 60(57-63) | 140 | <0.001 |
|  | >7 | 24(23.8-24.2) |  |  |
| 5th minute APGAR<br>score | <7 | 72(71-73) | 115 | <0.001 |
|  | >=7 | 30(25-35) |  |  |
| Perinatal asphyxia | Yes | 60(54-66) | 42.2 | <0.001 |
|  | No | 27(24-30) |  |  |
| Hemodynamic<br>instability | Yes | 70(64-76) | 51.6 | <0.001 |
|  | No | 30(25-35) |  |  |
| Weight for gestational<br>age | AFGA | 36(33-39) | 33.8 | <0.001 |
|  | SFGA | 72(72-73) |  |  |
| RDS | Yes | 70(68-72) | 87.2 | <0.001 |
|  | NO | 27(24-30) |  |  |
| Place of delivery | In born | 27(25-29) | 95.6 | <0.001 |
|  | Out born | 70(65-75) |  |  |
| Sepsis | Yes | 48(45-51) | 20.8 | <0.001 |
|  | No | 24(23.8-24.2) |  |  |
| Hypothermia | Yes | 48(41.1-58.9) | 11.5 | <0.001 |
|  | No | 24(22.7-25.2) |  |  |
| Kangaroo mother care<br>(KMC) | Yes | 24(23.7-24.3) | 132 | <0.001 |
|  | No | 65(61-69) |  |  |

### Predictors of Time to Initiate Trophic Feeding in Preterm Neonates

Findings from the bi-variate analysis showed that, BW,GA, first minute APGAR score, 5th minute APGAR score, hemodynamic instability, RDS, weight for gestational age, sepsis, hypothermia, Asphyxia, KMC, mode of delivery and place of delivery were candidate with time to initiate TF in preterm neonates admitted to NICU at P value < 0.25. Similarly in multivariate analysis, variables BW, first minute APGAR score, 5th minute APGAR score, RDS, and KMC service were remained as significant predictors of time to initiate TF among the preterm neonates admitted in NICU at P-value < 0.05.

**Table 4.**
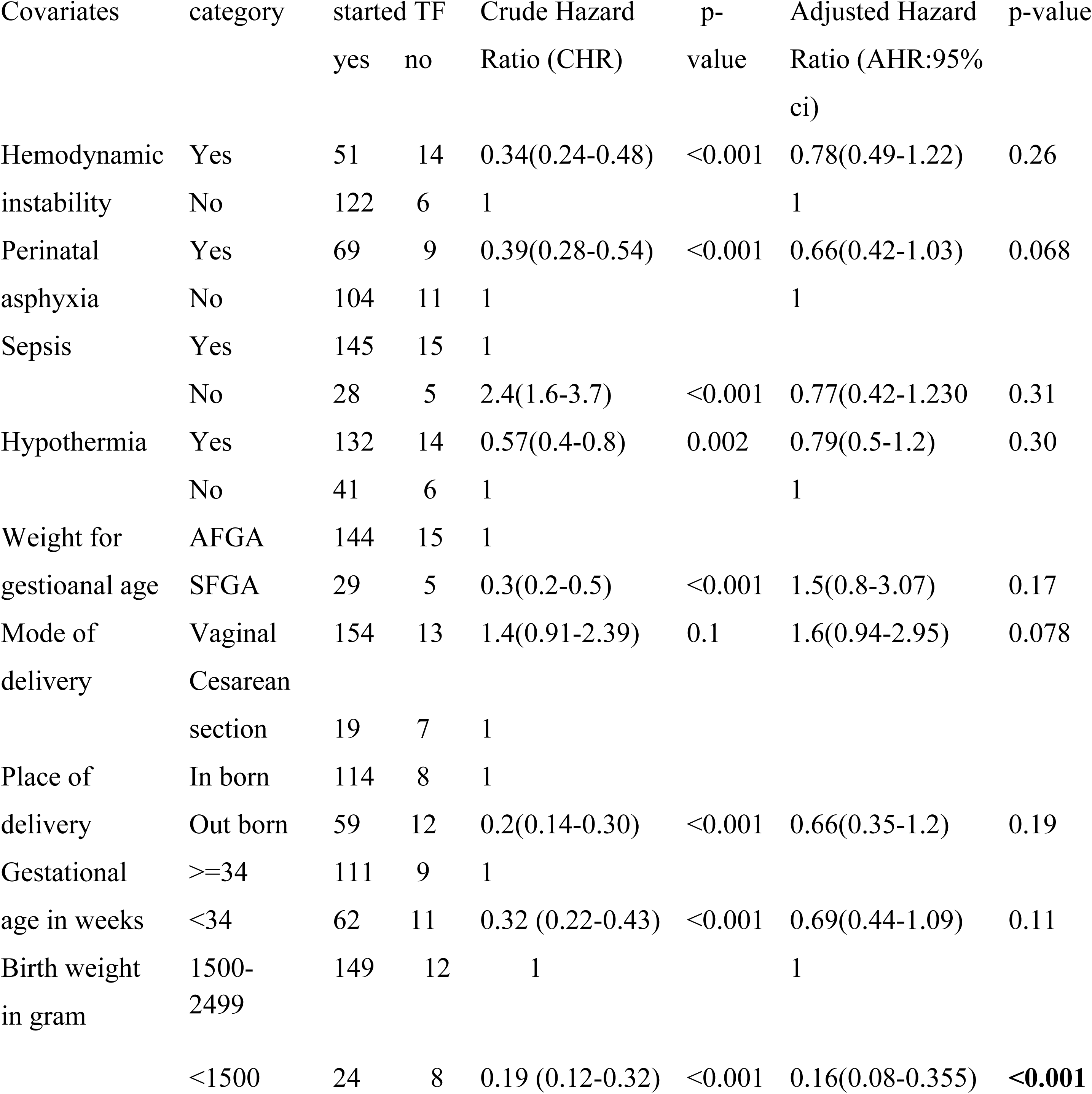
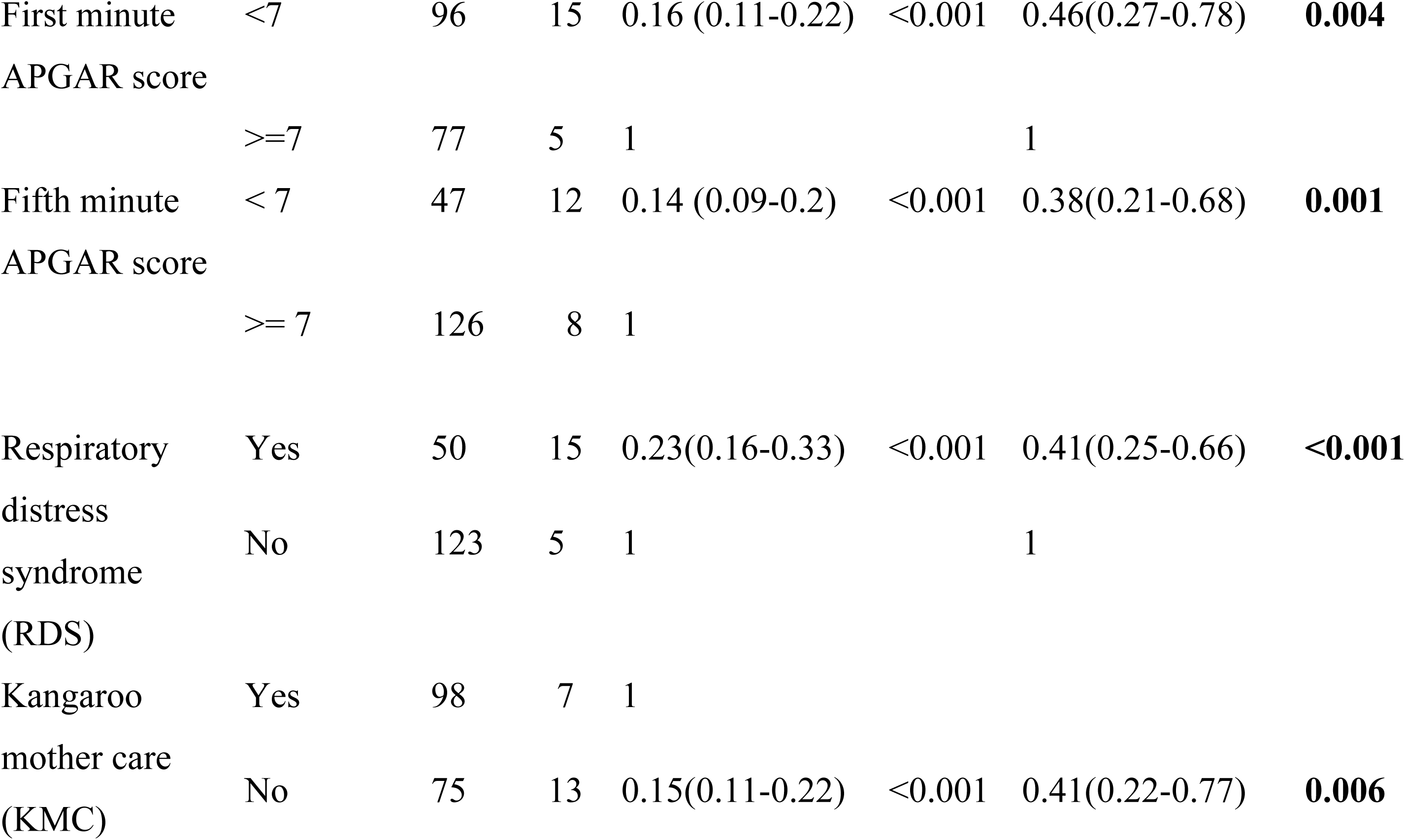
Results of bivariate and multivariate cox regression analysis of time to initiate TF and its predictor factor among preterm neonates admitted at public general Hospitals in NICU from December 20,2024 to February 30,2025,Tigray,Ethipia.(n=193).

The cox proportional hazard assumption was checked using overall global test for full model and it was (p=0.12).All covariates are met for proportional hazard assumption and residual was checked using the goodness of fit by cox Snell residuals.

In this study, the multivariate analysis revealed that the hazard of initiating TF among preterm neonates with a birth weight of less than 1500 grams was 84% lower compared to those born with a weight between 1500 and 2499 grams (AHR:0.16, 95% CI: 0.075-0.355). The hazard of initiating TF among preterm neonates with a first-minute APGAR score below seven was 54% lower compared to those with an APGAR score of seven and above (AHR: 0.46, 95% CI: 0.273-0.784). Similarly, the hazard of initiating TF among preterm neonates with a fifth-minute APGAR score below seven was 62% lower compared to those with a score of seven and above (AHR: 0.38, 95% CI: 0.211-0.679).

Additionally, the hazard of initiating TF was 59% lower among preterm neonates who had RDS compared to their counterparts (AHR: 0.41, 95% CI: 0.25-0.66).Finally, the hazard of initiating TF in the neonate without KMC service was associated with a 59% reduced likelihood of earlier TF initiation compared to the counterpart (AHR: 0.41, 95% CI:0.217-0.77).

## Discussion

This study aimed to assess the time to initiate TF and its predictors among preterm neonates admitted in the study hospital within the study period. In this study, the incidence of starting TF was 2 per 100 person hours of risk time. At the end of follow-up, 173(89.6%) of neonates (95% CI: 84.4–93.4) were started TF and 20 (10.4%) were censored.

This finding on the proportion of admitted preterm neonates who initiated TF is consistent with the study finding in, Addis Ababa 85%,(28). This may be due to similarity study design which is prospective cohort and sample size (193 vs153). However, it is higher than the studies done in southern Oromia (73.2%) (10). This discrepancy may be attributed to differences in the number of study areas and research design; prospective cohort studies facilitate real-time data collection, whereas retrospective cohort studies depend on pre-existing records and the number of study areas(31).

In this study 58 (30%) of preterm neonates started TF within the first 24 hours of birth, 58.5 % and 86% of preterm neonates started TF within 48 and 72 hours of birth, respectively. This indicates that only a small proportion of preterm neonates started TF within the first 24 hours of birth. However, this finding is lower than those reported in studies conducted in Nigeria and Kenya 48% and retrospective study in Portugal 44% observational study in Italy 74.1% (13, 32, 33).

The disparity in the initiation of TF among preterm neonates can be attributed to several factors, including healthcare infrastructure, provider knowledge on training and understanding of feeding protocols, and access to care, and socioeconomic conditions with better specialized facilities and resources, tend to show higher rates of early feeding (34).

In this prospective study, the median surviving time to initiate TF was 45 hours (95% CI: 42-56). This is similar study done southern Oromia 48 hours (10). but, higher than the finding reported in a prospective follow up study done in Addis Ababa 41 hours (28) and retrospective study done in North West Ethiopia 42 hours (14) the discrepancy might be regional healthcare practices, availability of specialized facilities, local policies regarding neonatal care, variability on providing TF feeding during the neonatal period and differences in healthcare infrastructure across regions.

In addition this result is lower than retrospective studies conducted in Portugal 72 hours (32).The difference might be due to sample size (193 vs 219),study period, study population, population difference and variation in neonatal management protocols (Ethiopia VS Portugal).

In this study, preterm neonates with a very low birth weight <1500gramwere 84% less likely to initiate TF on time compared to those with a BW greater than or equal to 1500gram with .This finding is higher than with the results of a retrospective follow-up study conducted in southern Oromia, Ethiopia 55% (10).This could be The conflict in Tigray has severely damaged health infrastructure, this destruction, combined with increased poverty and economic instability, Poor maternal nutrition and lack of healthcare access further exacerbate this issue, resulting in worse health outcomes compared to regions like Oromia, where healthcare resources are more available. Thus, the interplay of war, economic burden, and inadequate infrastructure significantly impacts the health of vulnerable newborns in Tigray (35, 36) and this could be underdevelopment of organs and lower readiness for enteral feeding in preterm neonates with lower birth weight (2).

Preterm neonates with an APGAR score below seven at one minute had a 54% lower likelihood of starting TF compared to those with a score of seven or higher. This finding is consistent with a retrospective study conducted in Northwest Ethiopia 40% (14) and prospective study in Addis Ababa 60% (28).

Similarly, the hazard of initiating TF among preterm neonates fifth minute APGAR score below seven was 62% less likely to start TF compared to neonates with APGAR score seven and above and this also supported by study conducted in Addis ababa,Ethipia 49% (28).This similarity might be low APGAR-score is the signal of intrauterine hypoxia which is an indication of decreased blood flow to gastrointestinal tract which might result in sever NEC that results in feeding intolerance (37).

The hazard of initiating TF preterm neonates with RDS had 59% less likely of initiating TF compared to those without the condition. This result aligns with a study conducted in the North West, which reported a 50% reduction (14) and Addis Ababa, Ethiopia (37%) (28). A possible explanation for this might be neonates with RDS often have rapid breathing, this may be due worsened by pressure in the abdominal, further compounded by physiological instability which heightens the risk of aspiration, feeding intolerance, and vomiting (38). So, this may influence professionals’ decision to delay starting TF. But, guidelines across the globe recommend a minimal TF can be initiated in neonates with RDS within the first 24 hours of life (39).

Finally, WHO recommends that KMC should be provided to preterm or LBW infants, beginning either in healthcare facilities or at home, and continuing for 8 to 24 hours each day (40). In accordance with this evidence, our study found that the absence of KMC was linked to a 59% lower likelihood of initiating TF early compared to those who received KMC.This finding is similar with a retrospective cohort study done in Oromia, Ethiopia 42% (10).This might be KMC improved survival rates, reduced infections, enhanced feeding and growth for the baby as well as bonding and maternal confidence (41).

### Limitations of the study

Demands regular monitoring and follow up

## Conclusion

This study identified a delay in the initiation of TF among preterm neonates compared with standard national guide lines, highlighting the need to enhance early feeding practices for this vulnerable group. Delays in TF initiation could impact neonatal health outcomes, such as increased risk of infections or prolonged hospital stays. Only 30% of the infants received TF within the first 24 hours, revealing a discrepancy between current practices and established guidelines. After adjusting for potential confounding factors, significant predictors of delayed TF initiation included very low birth weight, APGAR scores below seven at both one and five minutes, the presence of RDS, and neonate without KMC.

### Recommendations

**Healthcare providers**: should initiate TF as early as possible by addressing contributing factors and Provide specialized care and monitoring for high risk neonates with very low birth weight, low APGAR scores, and RDS to facilitate timely initiation of enteral feeding that may delay it by Adhering to national guidelines on the timely initiation of TF in preterm neonates to improve nutritional outcomes and reduce complications and Increase awareness and training on the importance and techniques of early trophic feeding, especially in NICU.

**Health facilities**: should create a safe and supportive environment to encourage and promote KMC practices for all eligible preterm neonates, recognizing its positive impact on the initiation of TF and Strength the neonatal care protocols that align with Ethiopian national guidelines for the initiation of trophic feeding and Provide continuous professional development and in-service training on neonatal nutrition practices for NICU staff.

Additionally, parents should be encouraged to actively participate in feeding plans and care decisions during their infant’s stay in the NICU.

**Ministry of health**: Enhance and develop early feeding protocols and implement standardized protocols to promote the initiation of enteral feeding within the first 24 hours of birth, especially for neonates with stable vital signs.

Finally, researchers should conduct long-term studies to assess the impact of early TF initiation on growth and neurodevelopmental outcomes in preterm infants in this area and to identify barriers to timely initiation of trophic feeding in various healthcare settings to guide to best practices for early TF initiation in preterm neonates and evaluate the impact of guideline adherence on short- and long-term outcomes in preterm neonates and explore effective interventions that can promote early initiation of trophic feeding in line with national practices.

## Declarations

### Ethical approval and consent to participant

An official ethical clearance letter was obtained from the Axum University College of Health Sciences Institutional Review Board (IRB) on 03 December 2024 with Protocol of Approval AKU-IRB 095/2024.Additionally, permission was secured from the Tigray Regional Health Bureau and the respective hospital medical director’s office. Upon receiving approval, communication was established with the medical directors, chief nursing directors, and intensive care unit heads at each General hospitals.

Following these steps, the designated personnel proceeded with following and obtaining the medical records of the selected samples. Throughout the process, greatest care was taken to ensure patient confidentiality. Given the prospective follow up nature of the study harm to individual patients was minimal as long as confidentiality was strictly maintained. To guarantee confidentiality, all collected data were anonymized using codes and stored securely in a locked room before being entered into a password-protected computer system. Patient names were excluded from the data collection format entirely. The principal investigator remained the sole individual with access to the data. All methods were performed in accordance with declarations of Helsinki.

### Consent for publication

Not applicable

### Availability of data and materials

The datasets used and/or analyzed during the current study are available

### Authors’ contribution

TW – developed the proposal, designed and participated in data collection, conducted the data analysis and interpretation, developed the first draft, revised subsequent drafts, and wrote the manuscript. ZB- participated on the conception of the research proposal, data analysis, and interpretation, reviewed and commented on successive drafts. AI- advised on the development of the research proposal, data analysis, and interpretation, reviewed and commented on successive drafts. BT – participated in the data collection, reviewing and editing the original draft. TG – participated in the data analysis, and data curation. NG- participated in the data analysis, and data curation. TM - participated in the reviewing and editing the original draft. ND - participated in the data analysis, and data curation .GG - participated in the data analysis, and data curation. All authors reviewed and approved the final draft of the manuscript and agreed to be personally accountable for the author’s own contributions.

### Authors’ information

TW (MSc. in Pediatric and child health nursing) clinical staff in Referral hospital Aksum University; ZB (MSc. in pediatric and child health nursing) Assistant Professor in Axum University; AI (MSc. in adult health Nursing) lecturer in Axum University; BT (MSc. in adult health Nursing) lecturer in Adigrat University; TG (MSc. in pediatric and child health nursing) lecturer in Araya Kahsu health science college; NG (MSc. in Pediatric and child health nursing clinical staff in Referral hospital Aksum University; TM (MPH.in epidemiology, MSc in adult health Nursing) lecturer in Axum University; ND (MSc. in pediatric and child health nursing) lecturer in Araya Kahsu health science college; GG (MSc. in pediatric and child health nursing) lecturer in Araya Kahsu health science college.

## Data Availability

All relevant data are within the manuscript and its Supporting Information files.

## Abbreviations

APGAR: Appearance, Pulse, Grammies, Activity, Respiration
BW: Birth weight
TF: Trophic Feeding
GA: Gestational Age
KMC: Kangaroo Mother Care
NEC: Necrotized Enterocolitis
NICU: Neonatal Intensive Care Unit
PNA: Perinatal Asphyxia
RDS: Respiratory Distress Syndrome
TF: Trophic feeding,

## Supporting information

S1 Dataset.

## ACKNOWLEDGMENT

I would like to express my sincere gratitude to my data collectors and supervisors for their facilitation, organization and collection of the data throughout the study period.

---

**Figure 1.**
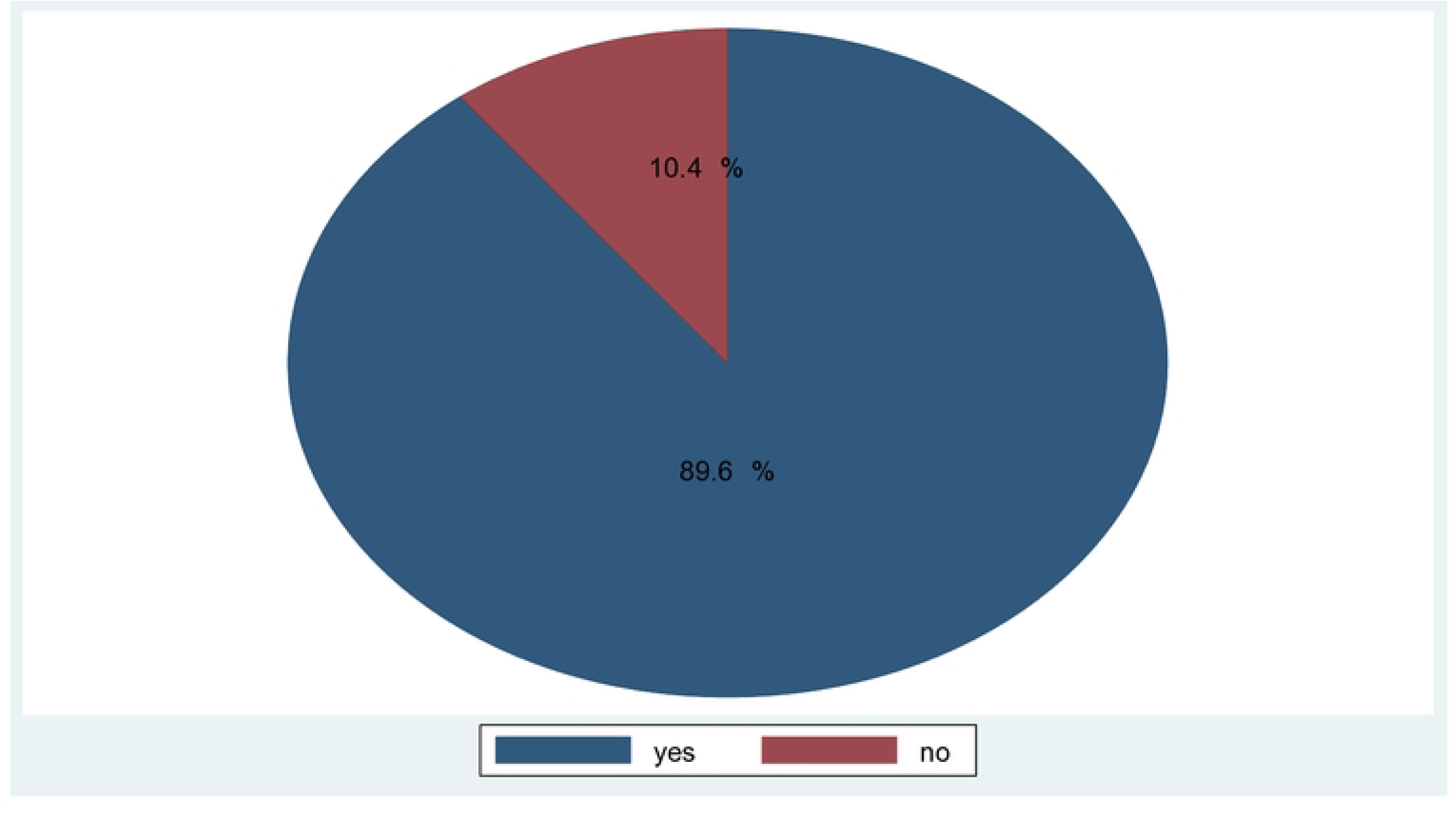
proportion of initiated trophic feeding and censored among preterm neonates admitted in NICU at General public Hospitals in Tigray, Ethiopia, 2024/25

**Figure 2.**
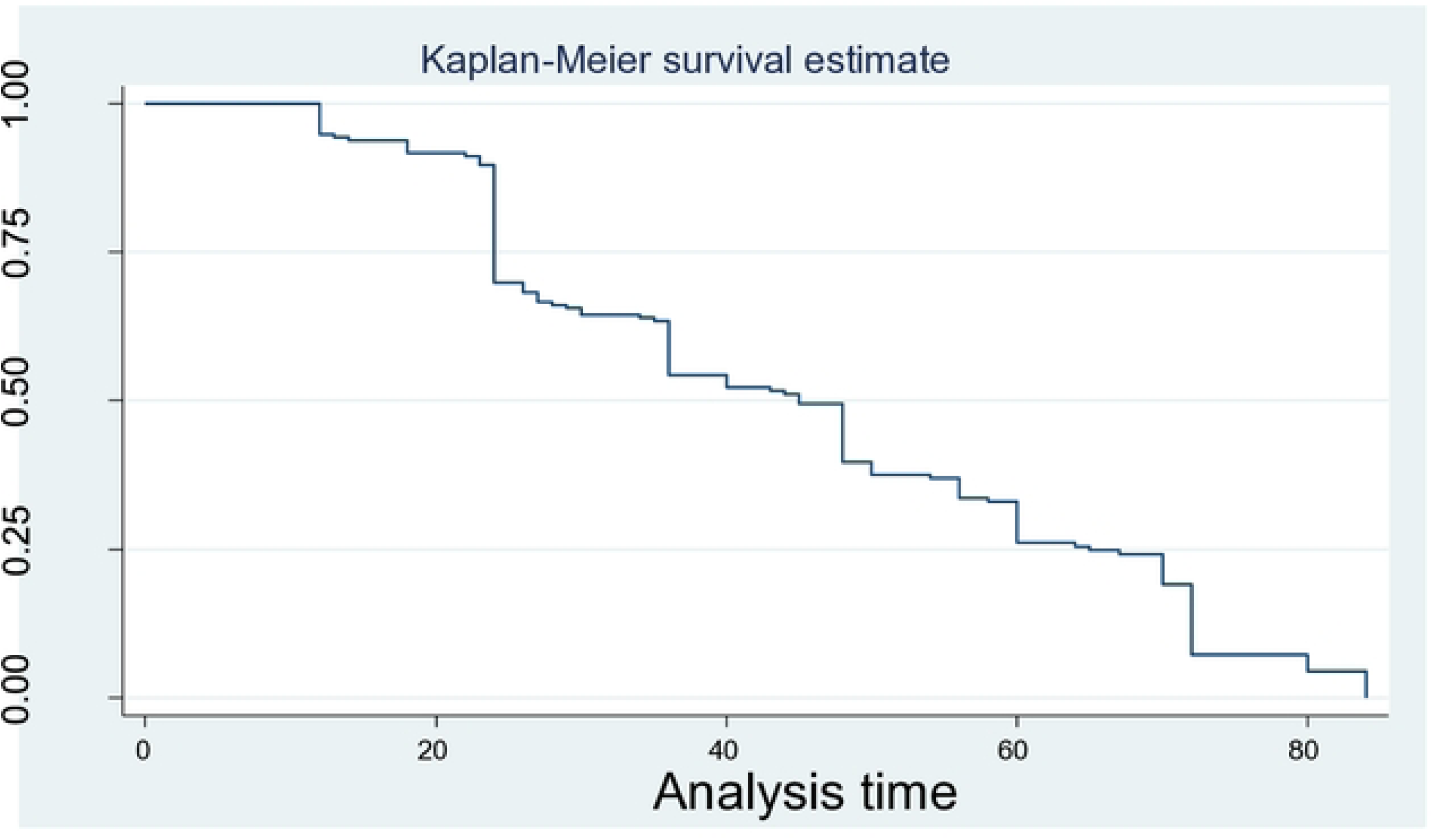
over all Kaplan Meier estimation time to initiate trophic feeding and its predictors among pretern neonates admitted in NICU in general public hospitals from December 20, 2024 to February 30, 2025, Tigray, Ethiopia.

**Figure 3.**
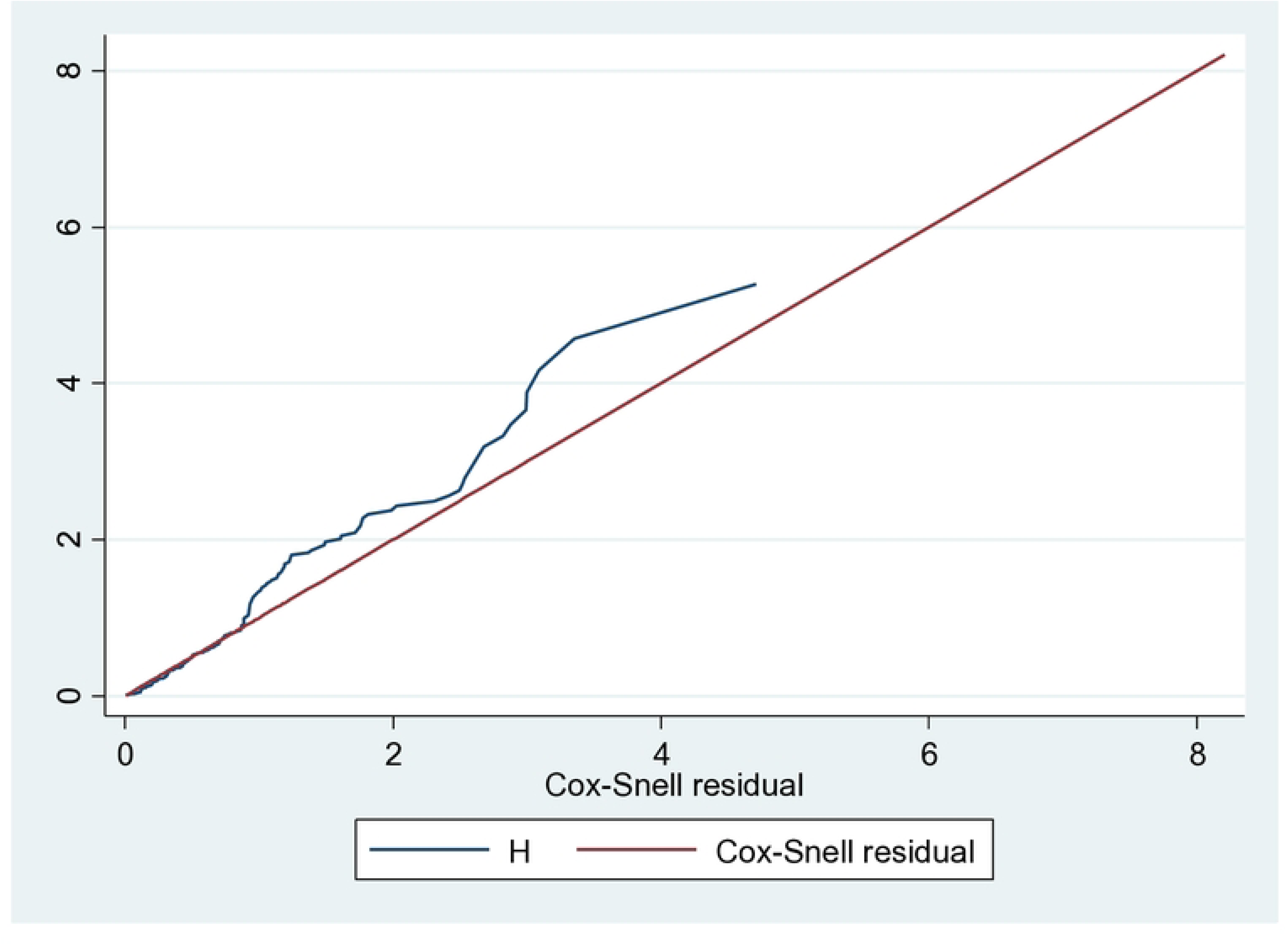
Nelson Aalen cumulative hazard graph against cox Snell residual on time to initiate trophic feeding and its predictors among pretern neonate admitted in public general hospitals from December 20, 2024 to February 30, 2025, Tigray Ethiopia.

**Figure 4.**
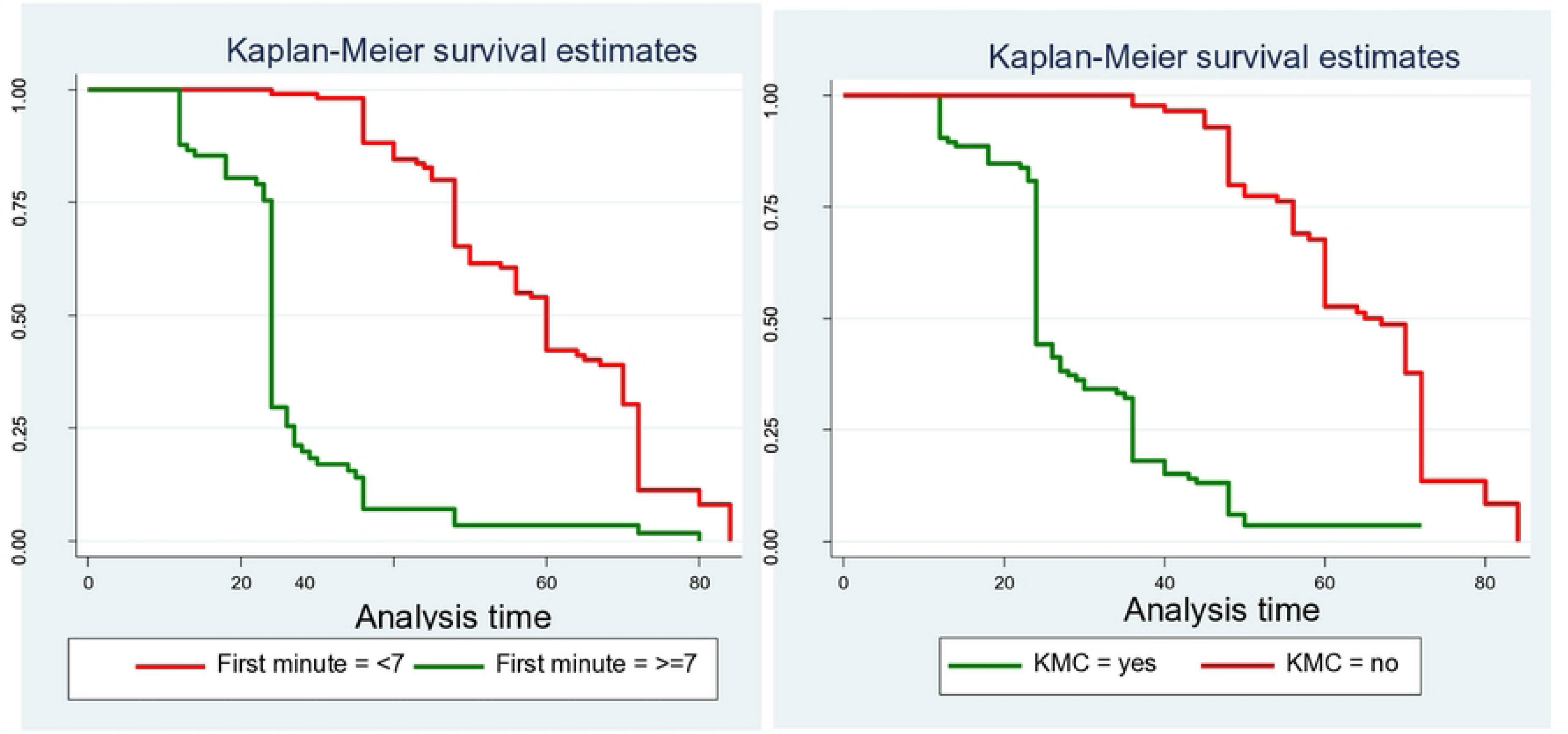
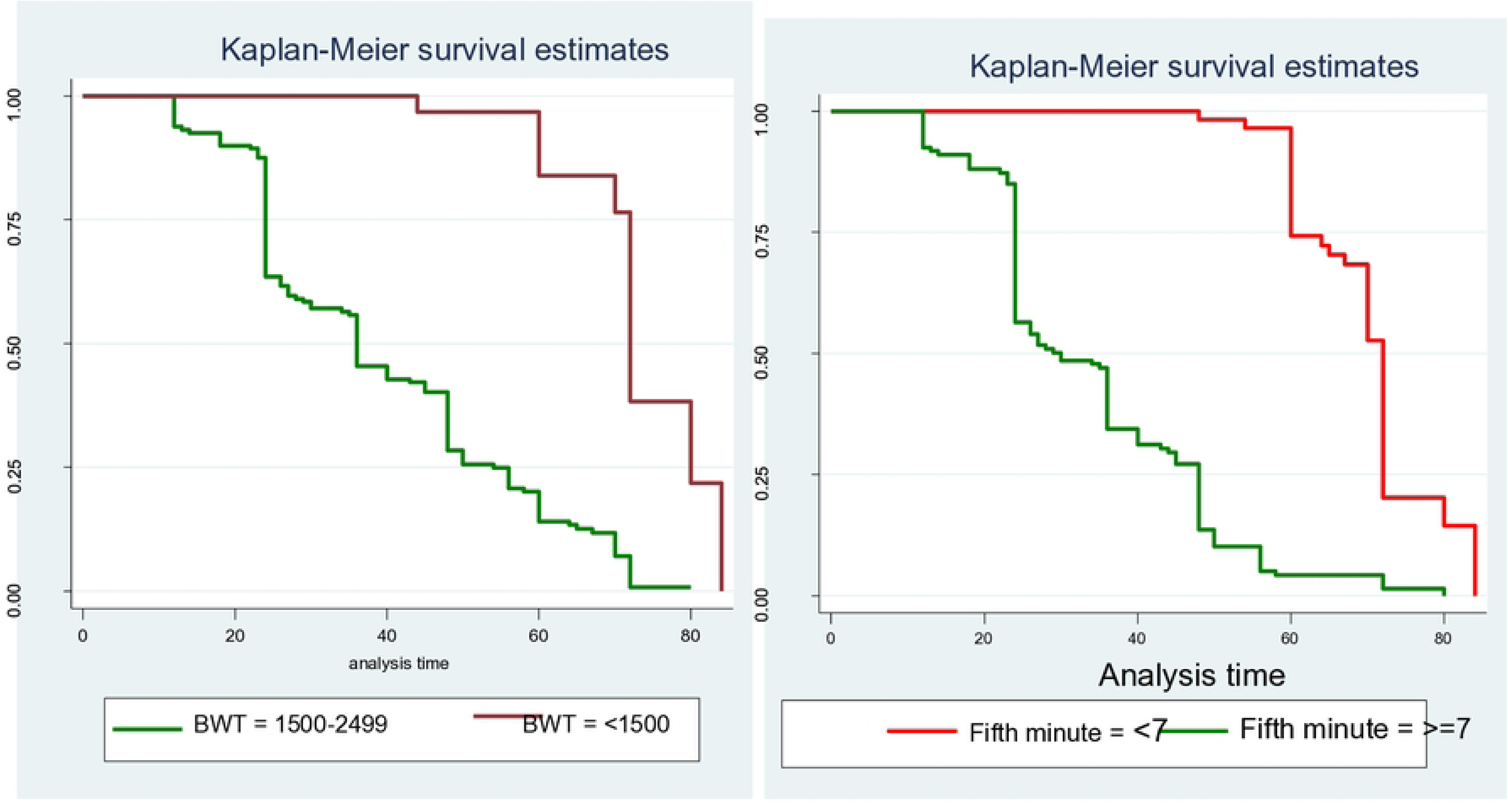
Log-Log plot of associated predictors

